# Duration of viral infectiousness and correlation with symptoms and diagnostic testing in non-hospitalized adults during acute SARS-CoV-2 infection: A longitudinal cohort study

**DOI:** 10.1101/2022.09.26.22280387

**Authors:** Paul K. Drain, Ronit R. Dalmat, Linhui Hao, Meagan J. Bemer, Elvira Budiawan, Jennifer F. Morton, Renee C. Ireton, Tien-Ying Hsiang, Zarna Marfatia, Roshni Prabhu, Claire Woosley, Adanech Gichamo, Elena Rechkina, Daphne Hamilton, Michalina Montaño, Jason L. Cantera, Alexey S. Ball, Inah Golez, Elise Smith, Alexander L. Greninger, M. Juliana McElrath, Matthew Thompson, Benjamin D. Grant, Allison Meisner, Geoffrey S. Gottlieb, Michael J. Gale

## Abstract

**Background:** Guidelines for SARS-CoV-2 have relied on limited data on duration of viral infectiousness and correlation with COVID-19 symptoms and diagnostic testing.

**Methods:** We enrolled ambulatory adults with acute SARS-CoV-2 infection and performed serial measurements of COVID-19 symptoms, nasal swab viral RNA, nucleocapsid (N) and spike (S) antigens, and replication-competent SARS-CoV-2 by culture. We determined average time from symptom onset to a first negative test result and estimated risk of infectiousness, as defined by a positive viral culture.

**Results:** Among 95 adults, median [interquartile range] time from symptom onset to first negative test result was 9 [5] days, 13 [6] days, 11 [4] days, and >19 days for S antigen, N antigen, viral culture growth, and viral RNA by RT-PCR, respectively. Beyond two weeks, viral cultures and N antigen titers were rarely positive, while viral RNA remained detectable among half (26/51) of participants tested 21-30 days after symptom onset. Between 6-10 days from symptom onset, N antigen was strongly associated with viral culture positivity (relative risk=7.61, 95% CI: 3.01-19.2), whereas neither viral RNA nor symptoms were associated with culture positivity. During the 14 days following symptom onset, presence of N antigen (adjusted relative risk=7.66, 95% CI: 3.96-14.82), remained strongly associated with viral culture positivity, regardless of COVID-19 symptoms.

**Conclusions:** Most adults have replication-competent SARS-CoV-2 for 10-14 after symptom onset, and N antigen testing is a strong predictor of viral infectiousness. Within two weeks from symptom onset, N antigen testing, rather than absence of symptoms or viral RNA, should be used to safely discontinue isolation.

**Funding:** Bill and Melinda Gates Foundation

## Introduction

Over 600 million cases of confirmed severe acute respiratory syndrome coronavirus-2 (SARS-CoV-2) infections and 6.5 million deaths from coronavirus disease (COVID-19) have been reported to the World Health Organization (WHO),^1^ and these numbers may be underestimated.^2,3^ Implementation of diagnostic testing for acute SARS-CoV-2 infection has been critical to identify COVID-19 cases, reduce transmission, and inform public health measures.^4^ Testing for SARS-CoV-2 relied on laboratory-based reverse transcriptase polymerase chain reaction (RT-PCR),^5^ but the emergence of rapid diagnostic tests has expanded equitable access to diagnostic testing worldwide.^6-8^

While antigen-based and nucleic acid amplification tests (NAATs) can diagnose SARS-CoV-2 infection and COVID-19 disease,^9-11^ the indication and interpretation of these tests may differ.^8^ Rapid antigen-based tests are considered less sensitive than NAATs,^12,13^ and have not been universally endorsed.^14^ However, rapid antigen-based tests have now become widely abundant in community settings,^15^ can detect emerging viral variants,^16^ and may be useful to facilitate testing and expedite treatment initiation.^8,17,18^

The presence of replication-competent virus, as measured by *in vitro* viral cultures, can serve as a proxy for individual infectivity or contagiousness,^19-22^ but, given the technical and biosafety resources required, routine testing by viral culture has not been feasible.^23,24^ As COVID-19 diagnostics and outpatient treatments become more accessible, there is a growing need to understand the duration of viral infectiousness and correlations with COVID-19 symptoms and diagnostic tests for acute, non-severe SARS-CoV-2 infection.^25^

Current public health guidance suggest a range of 5—20+ days on the duration of isolation for SARS-CoV-2-infected individuals to help reduce viral transmission.^26-28^ These recommendations depend on a person’s vaccination status, ongoing symptoms, and serial testing, but are inconsistent and informed by sparse data. Therefore, we characterized the kinetics and variations of viral RNA, viral antigens, and replication-competent virus, including isolation and viral growth assessment of several variants of interest/concern (VOI/VOC), during and after an acute SARS-CoV-2 infection to determine the duration of viral infectiousness with replication-competent virus, and predictors of ongoing individual infectiousness among COVID-19 symptoms and diagnostic tests for acute SARS-CoV-2 infection.

## Methods

### Study design and participants

We conducted a prospective cohort study with serial measurements among adults who had their first SARS-CoV-2 infection. Eligible participants were age >=18 years, had no known prior SARS-CoV-2 infection, and had not received COVID-19 vaccination. All SARS-CoV-2 infections were confirmed by RT-PCR from a nasal or nasopharyngeal swab within seven days of enrollment and participants did not require hospitalization. We excluded persons who were pregnant, had an immunological-altering condition (e.g., HIV, Type 1 diabetes mellitus, multiple sclerosis, lupus, and rheumatoid arthritis), were receiving immune-altering medications (e.g., glucocorticoids or immunomodulators), or were enrolled in an interventional COVID-19 clinical trial. The institutional review board at the University of Washington (STUDY00009981) approved the study.

### Procedures

Participants completed standardized questionnaires with comprehensive data, including onset and duration of COVID-19 symptoms.^29,30^ After enrollment, participants were scheduled for five additional clinical follow-up visits with pre-defined windows. During each clinical encounter, medical assistants obtained an anterior nasal (AN) swab (Puritan™ PurFlock™ Ultra Sterile Flocked Swabs 253806U), a nasopharyngeal (NP) swab (VWR Flocked Nasopharyngeal Specimen Swabs 97-2012), and venous blood. The NP swab was placed in viral transport medium (VTM) for SARS-CoV-2 and influenza A and B testing using RT-PCR on a Panther Fusion System (Hologic, Inc, Marlborough, USA). Residual VTM was stored at -80° C for viral cultures.

We isolated SARS-CoV-2 and assessed viral growth in culture regardless of RT-PCR result. We prepared two viral growth assays per sample using Vero E6 cells expressing human angiotensin-converting enzyme 2 and transmembrane Serine Protease 2 (VeroE6AT cells). We used microscopy to evaluate each inoculated well for syncytia formation and/or cellular death for 10 days. For positive viral cultures, we quantified virus titer as a median tissue culture infectious dose (TCID_50_) value using 10-fold serial dilutions. We performed whole genome viral sequencing from the culture isolates on an Illumina NextSeq 500 (Illumina, San Diego, USA), along with positive and negative controls. Sequence reads were processed, de-multiplexed, and assembled against the SARS-CoV-2 Wuhan-Hu-1 ancestral reference genome (NC_045512.2). For each genome, >1 million raw reads were acquired, representing >750x mean genome coverage and a minimum of 10x base coverage. Each consensus genome was analyzed and assigned a lineage based on Phylogenetic Assignment of Named Global Outbreak Lineages (Pangolin) nomenclature.^31^

We tested AN swabs for nucleocapsid (N) and spike (S) antigens using an electrochemiluminescence immunoassay.^32,33^ Dry AN swabs were resuspended in 500 µL VTM,^34^ incubated for 10 minutes at room temperature, and lysed with the addition of 1% Igepal CA-630. We added a heterophilic blocking reagent (Scantibodies, Santee, USA) to a concentration of 1.5 mg/mL to prevent non-specific binding. The N antigen assay used antibody pairs 40143-MM08 and 40143-MM05 (Sino Biological, Wayne, USA).^32,35^ The S antigen assay used antibody pairs 447 (AbCellera Biologics Inc., Vancouver, Canada) and 40591-MM43 (Sino Biological, Wayne, USA).^36^ Plates were read on a MESO QuickPlex SQ 120 plate reader (MesoScale Diagnostics, Rockville, USA) for quantitative concentrations. We fitted a four-parameter logistic function and calculated limits of detection (LOD).

We tested serum samples for SARS-CoV-2 total (IgG +IgM +IgA) anti-spike antibody titers using the Roche Cobas e411 system (Roche Molecular Diagnostics, Indianapolis, USA), and for SARS-CoV-2 anti-spike IgG antibody titers using a chemiluminescent microparticle immunoassay (Abbott Architect SARS-CoV-2 IgG II assay) on the AdviseDx platform (Abbott Diagnostics, Chicago, USA). The total anti-S antibody titers were reported as units per mL (U/mL), which were equivalent to a universal measurement of binding antibody units (BAU) per mL.^37^ The anti-S IgG antibody assay provides quantitative results with an “index value” being the ratio of the chemiluminescent signal between the test:calibration samples. Results were provided as arbitrary units per mL (AU/mL), which were then converted to binding antibody units (BAU) per mL [*antibody*(*BAU*/*mL*) = *antibody*(*AU*/*mL*)/7].^37^

### Statistical Analyses

We defined onset of symptoms as the day any COVID-19 symptom was appreciated by the participant. We calculated viral load from RT-PCR using a standard curve measuring Ct value of the Orf1 gene using the equation: *log*_10_ (*VL*) = -0.2909 * *Ct* + 12.953. We defined infectiousness as presence of any replication-competent SARS-CoV-2 in viral cultures, regardless of TCID_50_ value. Analyses of immunological responses were performed both including and excluding results from specimens collected after a COVID-19 vaccination. All analyses were conducted using R.

We used generalized estimating equations to estimate the relative risk of culture positivity for each diagnostic test. We stratified each model by the presence of a set of symptoms (three iterations of each test model): loss of taste/smell, fever, and respiratory symptoms. Additionally, we stratified results by categorical days since symptom onset (0-5, 6-10, 11-14 days), and limited analyses to visits with complete diagnostic testing. We performed separate analyses to estimate relative risk of culture positivity both overall and among people with symptoms, when adjusted for age, sex at birth, comorbidities, and variant.

We used LOESS to fit a smooth curve through the quantitative data corresponding to each testing modality by days from symptom onset. For plotting testing results below and above the limits of quantitation, we set values to one-half of the lower limit of quantitation or doubled the upper limit of quantitation, respectively. Median time from symptom onset to a first negative test result for viral antigen, replication-competent virus, and viral RNA was calculated among the individuals with a negative diagnostic test result during visits.

## Results

Among 106 recruited adults, four were ineligible and seven were excluded **(Suppl Figure 1)**. Among the 95 participants included, median age was 29 years, 43% were female, and most (67%) reported a known SARS-CoV-2 exposure **(Table 1)**. Median time from symptom onset to day of enrollment was six days **(Suppl Table 1)**. During the follow-up period, 30 (32%) participants received a COVID-19 vaccination. All participants tested negative for Influenza A and B.

**Table 1.** Characteristics of the study participants (N=95).

|  | **N (%)<sup>a</sup>** |
| --- | --- |
| Age (years); median [IQR] | 29 [24, 44] |
| Female sex assigned at birth | 41 (43) |
| Race and ethnicity |  |
| American Indian or Alaskan Native | 4 (4) |
| Native Hawaiian or Pacific Islander | 1 (1) |
| Hispanic or Latinx | 18 (19) |
| Asian | 11 (12) |
| White | 65 (68) |
| Black or African American | 8 (8) |
| Other race or ethnicity | 1 (1) |
| Comorbidities |  |
| Any chronic health condition | 16 (17) |
| Diabetes (Type II) | 2 (2) |
| Hypertension | 5 (5) |
| Other chronic health conditions<sup>b</sup> | 11 (12) |
| BMI (kg/m<sup>2</sup>); median [IQR] | 25.8 [22.9, 31.4] |
| SARS-CoV-2 exposure |  |
| Yes, known exposure | 64 (67) |
| If yes, exposure was within household | 46 (48) |
| No known exposure or don't know | 31 (33) |
| Symptoms at initial RT-PCR test date | 84 (88) |
| Days between positive RT-PCR test and enrollment; median [IQR] | 4 [3, 6] |
| Days between symptom onset and enrollment; median [IQR] | 6 [5, 8] |
| COVID-19-like symptoms in two weeks prior to enrollment | 95 (100) |
| Fatigue | 80 (84) |
| Cough | 70 (74) |
| Aches or muscle pains | 69 (73) |
| Headache | 69 (73) |
| Chills | 68 (72) |
| Runny nose | 60 (63) |
| Loss of taste | 53 (56) |
| Loss of smell | 53 (56) |
| Sore throat | 49 (52) |
| Fever | 46 (48) |
| Shortness of breath/difficulty breathing | 43 (45) |
| Diarrhea | 38 (40) |
| Nausea | 33 (35) |
| Vomiting | 10 (11) |
| Presence of symptoms at visit (n/N (%)) |  |
| Visit 4 (~14 days after enrollment) | 36/82 (44) |
| Visit 5 (~28 days after enrollment) | 3/79 (4)<sup>c</sup> |
| Visit 6 (~56 days after enrollment) | 5/79 (6)<sup>d</sup> |
| COVID-19 Vaccination Status |  |
| Vaccinated within 1 month of enrollment | 8 (8) |
| Vaccinated within 2 months of enrollment | 22 (23) |
| Vaccinated during study period | 30 (32) |
| SARS-CoV-2 Variant |  |
| Alpha | 9 (9) |
| Epsilon | 22 (23) |
| Gamma | 1 (1) |
| Other (non-VOI/VOC) | 28 (29) |
| No known Pangolin lineage | 2 (2) |
| Not sequenced | 33 (35) |

BMI=body mass index; IQR=interquartile range; VOI=variant of interest; VOC=variant of concern.
<sup>a</sup>Percentages may not add to 100 due to rounding.
<sup>b</sup>Other included lung conditions (n=5, 2 with asthma and 1 with COPD), hypothyroidism (n=1), and not otherwise specified (n=5). No participants reported other chronic heart, kidney, or liver conditions.
<sup>c</sup>3 of 3 participants reporting symptoms at V5 reported symptoms at V4; 0 of 3 did not attend V4.
<sup>d</sup>1 of 5 participants reporting symptoms at V6 reported symptoms at V5; 1 of 5 did not attend V5. Of the 3 participants who were symptomatic at V6 and asymptomatic at V5, 3 reported symptoms at V4. At V6, the symptoms reported by these 3 participants were loss of sense of taste and loss of sense of smell.

Sixty participants had a sequenced viral variant identified using Pango lineage designations: 32 had VOI/VOC virus (9 alpha, 22 epsilon, 1 gamma) and 28 had non-VOI/VOC virus **(Table 1)**. Two participants known to be household contacts had related viral isolates that did not match a known lineage. Phylogenetic analyses indicated a diversity of viral lineages compared to known reference strains **(Figure 1)**. In stratified analyses, there were no major differences in cohort characteristics or disease presentation by variant.

**Figure 1.**
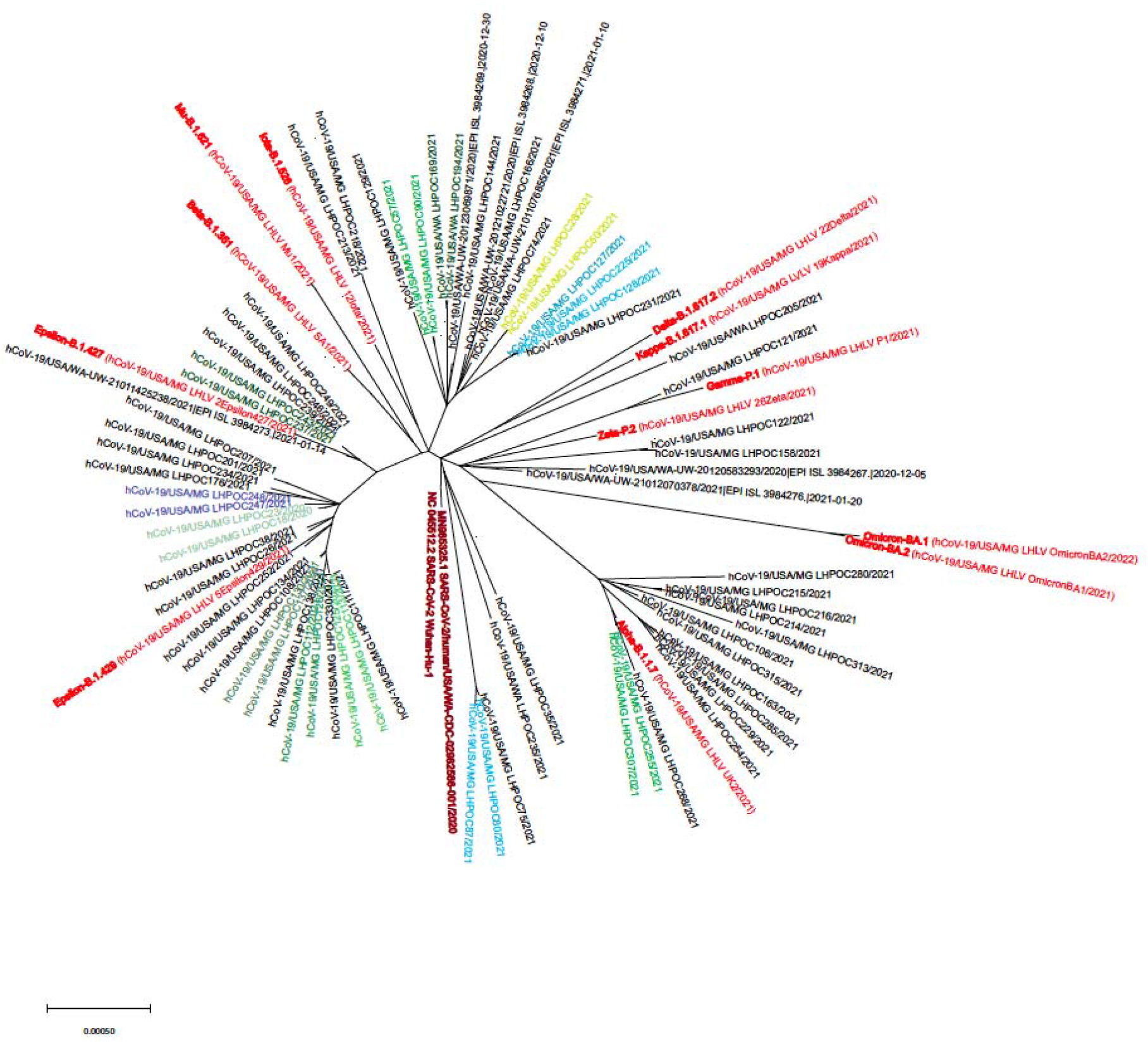
Viral phylogram for unvaccinated adults presenting with acute SARS-CoV-2 infection. Phylogram on the SARS-CoV-2 virus and variants among the enrolled participants (red font indicates reference strains; similar colors indicate related samples, either household contacts or samples from the same individual at different time points).

When categorizing participants by RT-PCR, viral culture, and N antigen positivity, the vast majority (80%) were positive by all three measures during the first 5 days from symptom onset **(Table 2)**. Between 6-10 days from symptom onset, most (96%; 68/71) samples were positive by RT-PCR, 79% (56/71) were N antigen-positive, and 41% (29/71) were culture-positive. Between 11-15 days from symptom onset, 8% (8/96) tests were culture positive and 6% (6/96) were negative by all three tests. Most were positive by RT-PCR and negative by culture (85%; 82/96). Of these 82 tests, 39% were antigen positive and 61% were antigen negative. Beyond 15 days, viral cultures and N antigen titers were rarely positive. Conversely, the RT-PCR test remained positive in 60% (62/104) of participants between 16-30 days after onset of symptoms. Among those, 51% (26/51) of participants remained positive by RT-PCR test between 21-30 days after symptom onset. Overall, the estimated median [interquartile range] time from symptom onset to first negative test result was 9 [5] days, 11 [4] days, 13 [6] days, and >19 days for S antigen, viral culture growth, N antigen, and viral RNA by RT-PCR, respectively **(Figure 2, Suppl Table 2)**.

**Table 2.** Diagnostic test kinetics of RT-PCR, culture, and nucleocapsid (N) antigen positivity, categorized by days since symptom onset.

| Measure |  |  |  | Days since symptom onset<sup>a</sup> |  |  |  |  |  | Total |
| --- | --- | --- | --- | --- | --- | --- | --- | --- | --- | --- |
| RT-PCR | Culture | N antigen | --- | 0-5<br>% (n) | 6-10<br>% (n) | 11-15<br>% (n) | 16-20<br>% (n) | 21-25<br>% (n) | 26-30<br>% (n) | % (N) |
| + | + | + | All positive | 80 (28) | 41 (29) | 1 (1) | 0 (0) | 0 (0) | 0 (0) | 19 (58) |
| + | + | - | RT-PCR/culture positive | 3 (1) | 0 (0) | 5 (5) | 4 (2) | 0 (0) | 8 (1) | 3 (9) |
| + | - | + | RT-PCR/antigen positive | 6 (2) | 37 (26) | 33 (32) | 6 (3) | 3 (1) | 0 (0) | 21 (64) |
| + | - | - | Only RT-PCR positive | 3 (1) | 18 (13) | 52 (50) | 58 (31) | 46 (18) | 50 (6) | 39 (119) |
| - | + | + | Culture/antigen positive | 0 (0) | 0 (0) | 0 (0) | 0 (0) | 0 (0) | 0 (0) | 0 (0) |
| - | + | - | Only culture positive | 0 (0) | 0 (0) | 2 (2) | 0 (0) | 0 (0) | 0 (0) | 1 (2) |
| - | - | + | Only antigen positive | 0 (0) | 1 (1) | 0 (0) | 2 (1) | 3 (1) | 0 (0) | 1 (3) |
| - | - | - | All negative | 9 (3) | 3 (2) | 6 (6) | 30 (16) | 49 (19) | 42 (5) | 17 (51) |
| Column total |  |  |  | 35 | 71 | 96 | 53 | 39 | 12 | 306 |

<sup>a</sup>Color shading indicates the relative percentage of people within the 5-day periods of days since symptom onset, ranging from dark green ( $\geq 80\%$ ) to no shading (0%).

**Figure 2.**
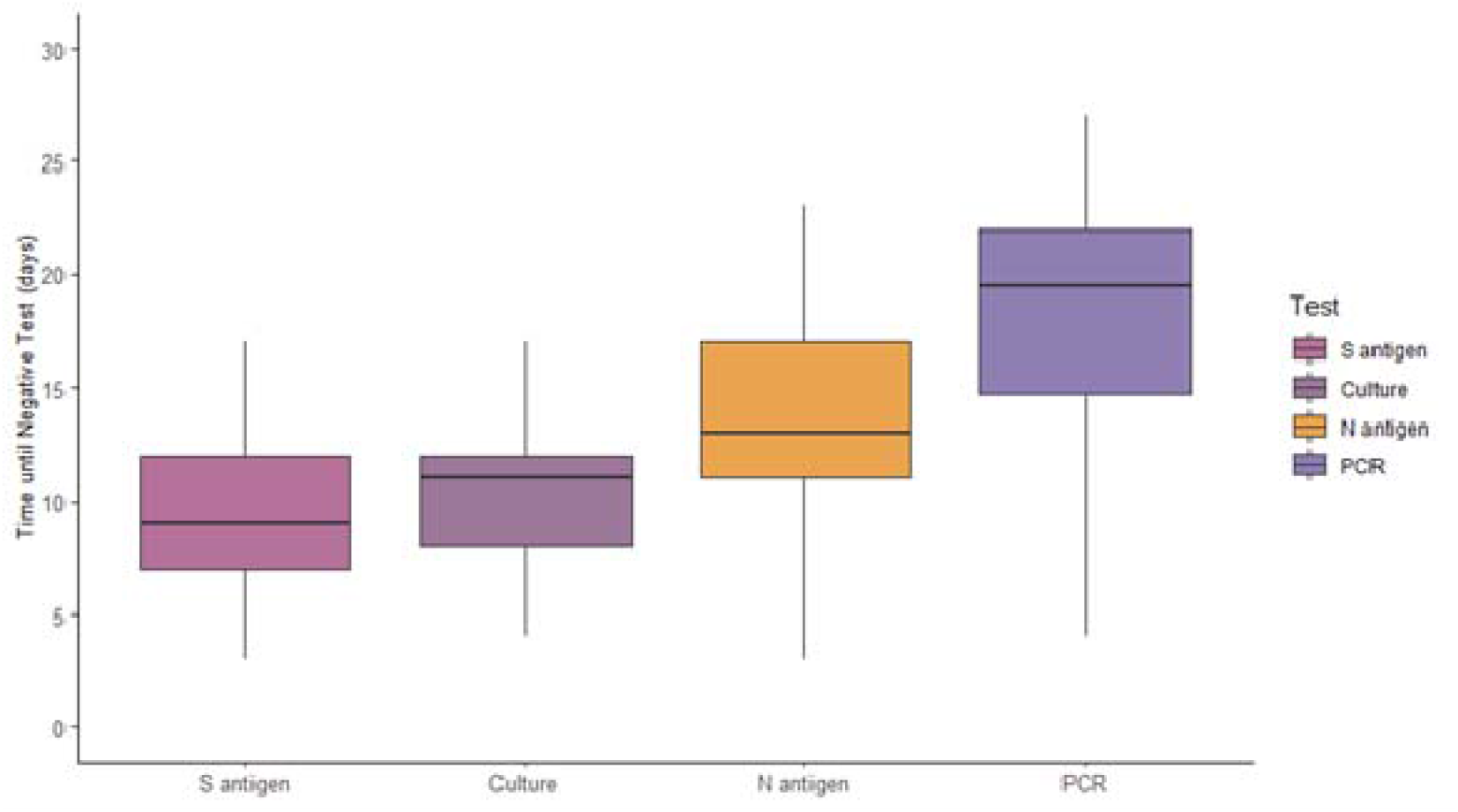
Median days from symptom onset to first negative test among spike (S) antigen, viral culture, nucleocapsid (N) antigen, and RT-PCR for viral RNA. Median [interquartile range] days from symptom onset to the first negative test was 9 [5] days for S antigen, 11 [4] days for viral culture, 13 [6] days for N antigen, and >19 days for RT-PCR, among participants testing negative within 14 days of enrollment. Median for RT-PCR could not be precisely approximated because more than half of participants were positive at all available sample times (within 14 days of enrollment, which corresponds to 2-4 weeks after onset of symptoms).

We observed a few instances (n=4) of apparent viral “rebound” where Ct values were above Ct >30 but then declined to ≤30 on a subsequent test **(Suppl Figure 3A)**. In two occurrences where the rebound was documented from an RT-PCR test collected more than ten days after symptom onset, a culture of the residual VTM, as well as an N antigen test from an anterior nares swab on the same day, were both negative. One participant displayed an apparent viral “rebound” on two separate occurrences. For the one occurrence at nine days since symptom onset, both the culture and N antigen test result were positive. A second “rebound” occurrence at 19 days since symptom onset, a culture of the residual VTM was positive and N antigen test was negative.

Across the cohort, presence of fever, respiratory symptoms, and loss of taste/smell were not statistically significantly associated with infectiousness during the first 14 days after onset of symptoms **(Table 3)**. Between 6-10 days from symptom onset, presence of N antigen was significantly associated with viral culture positivity [relative risk (RR)=7.61, 95% CI: 3.01-19.22], whereas presence of viral RNA was not statistically significantly associated with culture positivity. Presence of N antigen remained strongly associated with greater risk of infectiousness, despite presence/absence of COVID-19 symptoms. When adjusted for age, sex, comorbidities, and viral variant, presence of N antigen was strongly associated with higher risk of infectiousness within two weeks from symptom onset, both overall and among those with symptoms (overall aRR=7.66, 95% CI: 3.96-14.82) **(Table 4)**. In similar adjusted analyses, RT-PCR positivity was not statistically significantly associated with risk of infectiousness when adjusted for symptoms, but the association was statistically significant among those with fever (aRR=4.12, 95% CI: 1.06-15.91) or respiratory symptoms (aRR=4.25, 95% CI: 1.15-15.66).

**Table 3.**
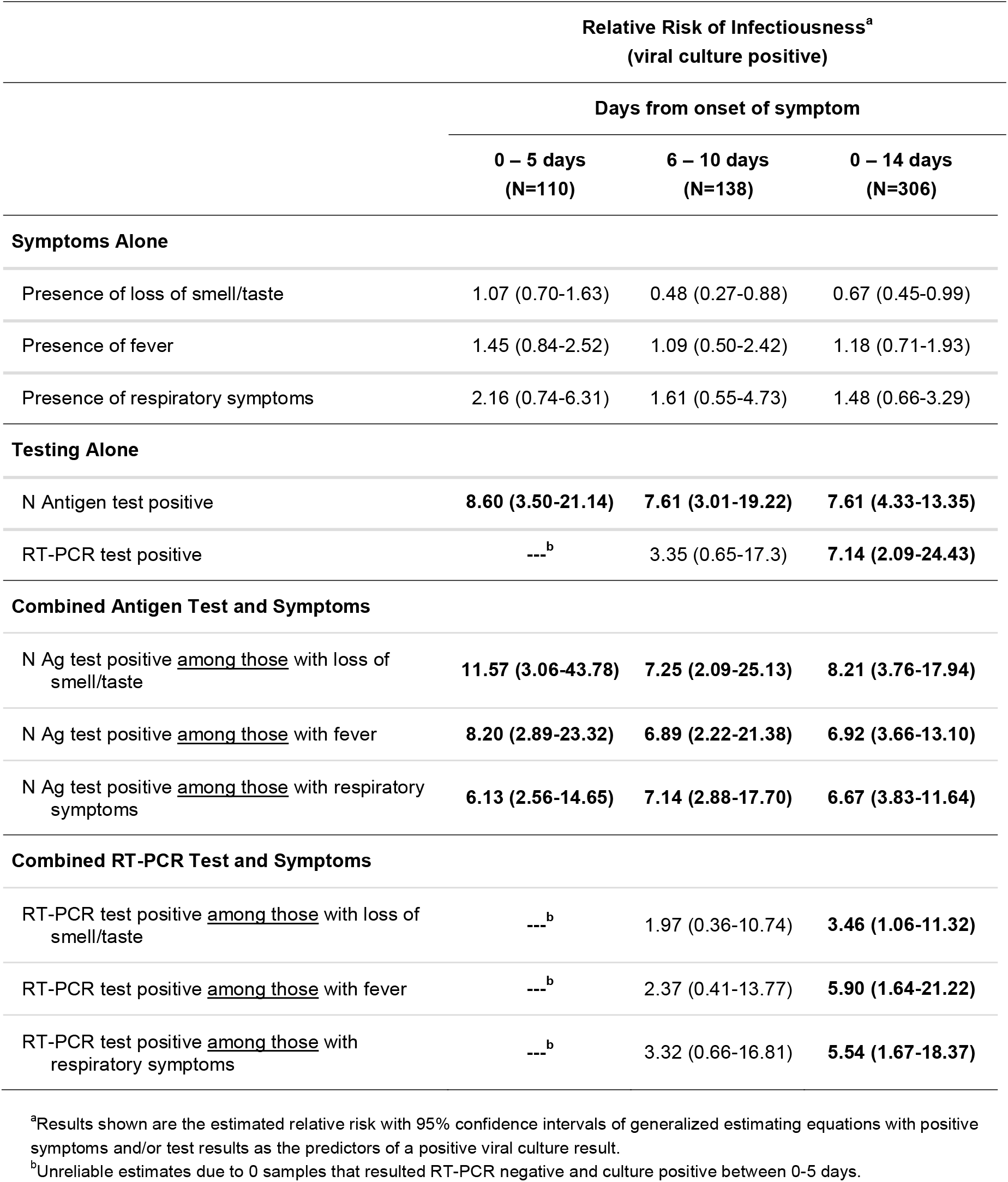
Estimates of relative risk of infectiousness (viral culture positive) based on symptoms (loss of taste/smell, fever, or respiratory), nucleocapsid (N) antigen or RT-PCR test result, and combinations, stratified by days since symptom onset.

**Table 4.** Estimates of adjusted relative risk of infectiousness (viral culture positive) based on nucleocapsid (N) antigen or RT-PCR test result between 0-14 days since symptom onset, and stratified by symptoms (loss of taste/smell, fever, or respiratory).

|  | **Adjusted Relative Risk of Infectiousness<sup>a</sup>** |  |  |  |
| --- | --- | --- | --- | --- |
|  | **(viral culture positive)** |  |  |  |
|  | **Regardless of**<br>**symptoms** | **Persons with**<br>**loss of smell/taste** | **Persons with**<br>**fever** | **Persons with**<br>**respiratory symptoms** |
|  | **(N=306)** | **(N=198)** | **(N=226)** | **(N=268)** |
| N Antigen test positive | **7.66**<br>**(3.96-14.82)** | **7.33**<br>**(3.30-16.32)** | **4.77**<br>**(2.56-8.89)** | **5.11**<br>**(2.92-8.94)** |
| RT-PCR test positive | 2.74<br>(0.81-9.25) | 2.57<br>(0.75-8.79) | **4.12**<br>**(1.06-15.91)** | **4.25**<br>**(1.15-15.66)** |

<sup>a</sup>Results shown are the estimated relative risk with 95% confidence intervals of generalized estimating equations with positive symptoms and/or test results as the predictors of a positive viral culture result. All models were adjusted for age, sex at birth, comorbidities, and SARS-CoV-2 variant.

We used LOESS curves to describe the clinical and diagnostic trajectories by days since symptom onset **(Figure 3)**. Most participants reported COVID-19 symptoms through 14 days. Replication-competent virus was routinely present in NP swabs through seven days, while only two participants were culture-positive beyond 15 days from symptom onset. One unique individual had trace viral growth (TCID_50_ <100) at 26 days since symptom onset, which was sequenced as viral lineage B.1.1.7. At 19 days post-symptom onset, this participant was RT-PCR positive (Ct=39.0), viral culture negative, and N antigen negative. Seven days later, the same participant remained RT-PCR positive (Ct=38.3), N antigen negative, and asymptomatic.

**Figure 3.**
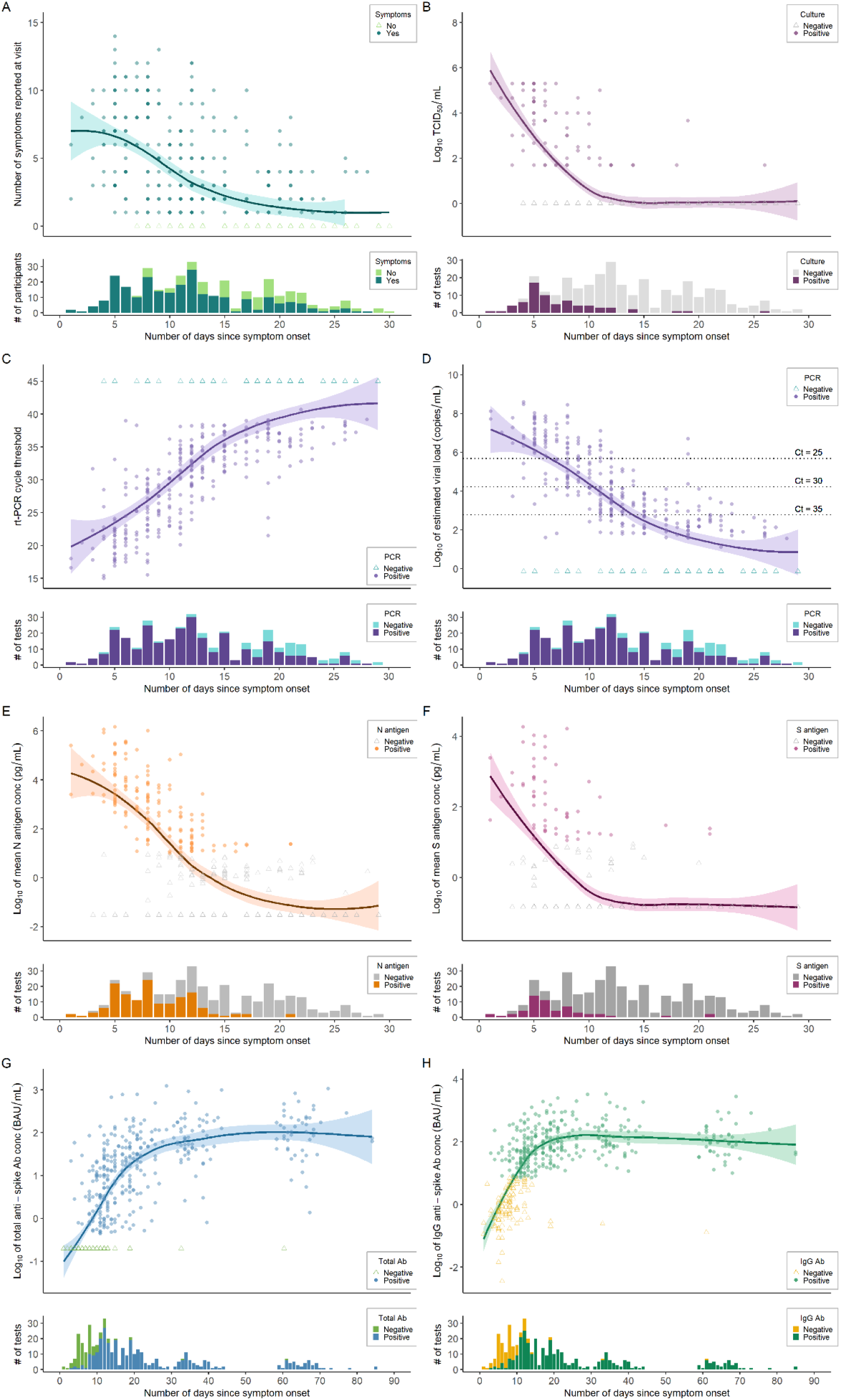
Trajectory of clinical symptoms, replication-competent viral growth, viral load by RT-PCR, nucleocapsid and spike antigen concentrations, and antibody titers, by days since symptom onset. Within each panel, quantitative data are displayed in the top portion and dichotomous positive/negative test results are displayed in the bottom portion, by number of days since date of symptom onset. Average lines represent LOESS curves and shaded regions represent 95% confidence intervals. Darker coloring indicates higher density of observations at that value (e.g., overlapping points lead to darker coloration). **A**, Total number of COVID-19 symptoms reported by participants at each clinical visit (n=351). **B**, Viral culture by log_10_ TCID_50_ per mL, with an estimated limit of detection of 2.0 (n=307). **C**, RT-PCR testing of viral RNA with cycle threshold (Ct) on the vertical axis (n=347). **D**, Estimated log_10_ of SARS-CoV-2 viral load (copies/mL) from RT-PCR testing (n=347). **E**, Nucleocapsid (N) antigen log_10_ mean concentration (pg/mL) as measured by a MesoScale Diagnostics assay (n=348). **F**, Spike (S) antigen log_10_ mean concentration (pg/mL) as measured by a MesoScale Diagnostics assay (n=349). **G**, Total anti-spike log_10_ mean antibody concentration (BAU/mL) tested by Roche Elecsys assay (n=442), excluding antibody titers obtained after vaccination. **H**, Anti-spike IgG log_10_ mean antibody concentration (BAU/mL) tested by Abbott AdviseDx assay (n=442), excluding antibody titers obtained after vaccination.

The average peak viral load was 6-8 log_10_ copies/mL (Ct=16-24), which declined to an average viral load of 2-3 log_10_ copies/mL (Ct≈35) after 14 days from symptom onset. While most people remained RT-PCR positive after 14 days, only 3 participants had a Ct <30 cycles beyond 14 days. Antigenic titers of N and S proteins had lower relative concentrations and faster rate of decline. After 14 days, the vast majority of participants tested negative for both N and S antigens. Humoral immune responses, as measured by total and IgG anti-spike antibodies, appeared within 14 days of symptom onset, plateaued within 30 days, and remained durable between 60-70 days. When including samples collected after COVID-19 vaccination, both total and IgG anti-spike antibody titers were appreciably higher and with a more durable trajectory **(Suppl Figure 2)**. We aggregated the findings of diagnostic test kinetics, infectivity, and immunological responses to illustrate the empirical trajectory of results during acute SARS-CoV-2 infection **(Figure 4)**.

**Figure 4.**
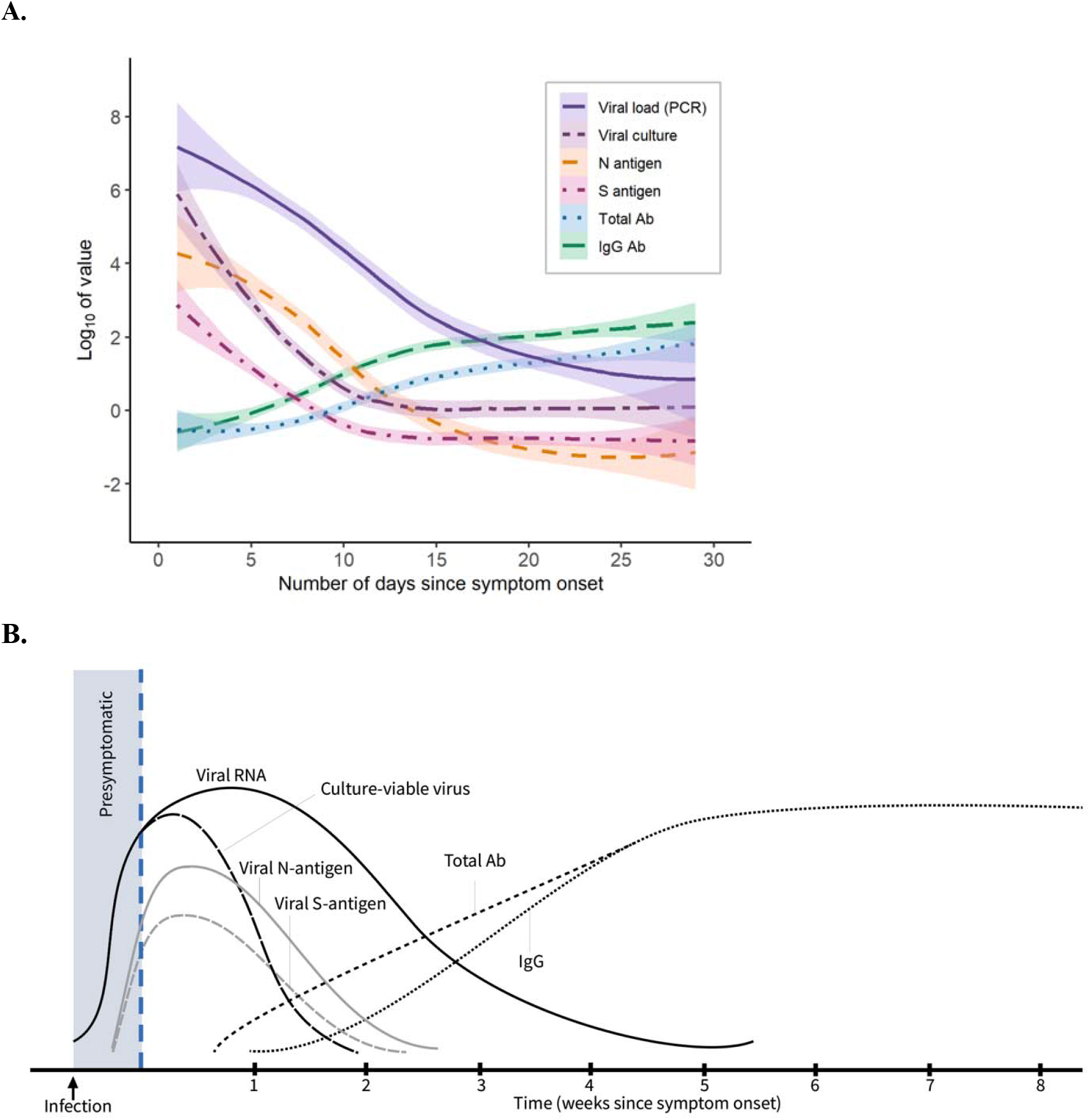
Diagnostic test kinetics and immunological responses in adults with non-severe, symptomatic SARS-CoV-2 infection. Average lines represent LOESS curves and shaded regions represent 95% confidence intervals. The x-axis shows days since symptom onset and the y-axis uses a log transformation. **A**, Study data with Log_10_ values for measured SARS-CoV-2 viral load, TCID_50_ from viral culture, nucleocapsid (N) antigen and spike (S) antigen mean concentration, and total anti-spike and anti-spike IgG antibody concentrations, by number of days since symptom onset. **B**, Theoretical model of diagnostic test kinetics and immunological responses, as extrapolated from observed data obtained among unvaccinated adults during acute SARS-CoV-2 infection.

## Discussion

Among ambulatory adults with community-acquired SARS-CoV-2 infection, the period of infectiousness averaged 11 days after onset of symptoms and extended to 15 days for several individuals. During this time period, N antigen testing was the strongest predictor of the risk of infectiousness and was superior to COVID-19 symptom monitoring and molecular testing. RT-PCR and N antigen assay results correlated with viral culture within the first 5 days from symptom onset, while N antigen test results remained significantly associated with infectiousness during the 6-10 day window and across the 0-14 day period after onset of symptoms. Overall, molecular testing by RT-PCR was a highly sensitive test for initial diagnosis, and presence of N antigen in nasal swabs was optimal for determining the potential infectiousness during the subsequent isolation period.

Most participants had high SARS-CoV-2 viral load (Ct<25), N antigen positivity, and a positive viral culture within 7 days of symptom onset. While most participants’ viral load measurements decreased steadily during the subsequent two weeks of follow-up, a few participants experienced a late viral rebound. Separate plots for persons having initial N antigen positivity, RT-PCR positivity, and viral isolation/culture positivity—all within five days of symptom onset—demonstrated considerable individual heterogeneity of diagnostic test results and trajectories. In our cohort, one participant with prolonged viral growth was unlikely to have had a second infection, based on phylogenetic analyses, but had prolonged low levels of replication-competent virus.

Prior models of diagnostic test kinetics, including our own, have relied on limited studies to generate theoretical models.^8,38^ Generating empiric longitudinal measurements of viral burden, viral sequence, and immunological responses helped identify the temporal diagnostic test kinetics and heterogeneity among infection-naïve individuals infected with SARS-CoV-2 virus. RT-PCR testing has high diagnostic sensitivity and utility for surveillance of emerging viral variants. Lower relative concentrations of both N and S antigenic titers may contribute to a lower overall diagnostic sensitivity of rapid antigen tests, when compared to RT-PCR. However, the rate of decline for N antigen level tracks more closely with the decrease in replication-competent virus in NP swabs, which may serve as a proxy for potential infectivity.

Other studies have also reported prolonged positive RT-PCR test results for 1-3 months after initial infection.^19,39-43^ In our cohort, positive viral cultures mostly occurred in samples with a high viral load (Ct <35). However, low levels of replication-competent virus (below limit of precise TCID_50_ quantification) were isolated in four specimens with a low or undetectable viral loads (one with Ct=35.3; one with Ct=38.3; two were RT-PCR negative) and N antigen test negative. Therefore, our data also suggest that positive RT-PCR specimens with a low viral load (Ct value >≈35 cycles) are unlikely to correlate with recovery of substantial amounts of replication-competent SARS-CoV-2 virus. While TCID_50_ has itself not been empirically linked to risk of transmission, our results indicate that TCID_50_ may serve as a measure of the presence of transmissible virus and NAATs may detect remnant viral RNA beyond the window of infectivity.^44-46^

The genetic diversity of cultured virus within our cohort reflects the local and temporal viral dynamics that occurred during our study period. We observed little viral genetic variation within individual subjects across study time points. Among ten individuals with serial virus isolation and three household contact groups, we observed 0-19 and 0-14 nucleotides changes between viral isolates, respectively. Therefore, the viral genome remained highly conserved over the study time course and across likely transmission events. Thus far, several studies have evaluated the longitudinal diagnostic kinetics among hospitalized adults,^47-50^ but up to now no studies have evaluated synchronous changes in the longitudinal biomarkers, diagnostic kinetics, infectiousness, and immunological responses among ambulatory adults with acute non-severe SARS-CoV-2 infection.

These results, when combined in a comprehensive diagnostic model of acute SARS-CoV-2 infection may help inform testing guidelines and public health practice. The estimated relative risk of culture positivity for a positive N antigen test (versus a negative N antigen test) was robust, regardless of presence of fever or time since symptom onset within 14 days. Therefore, for public health practices, N antigen testing may be the preferred method of testing to determine the recovery of replication-competent virus and potential infectivity between 6-10 days (or 0-14 days) from symptom onset, either with or without the presence of symptoms. Since persons with symptomatic, non-severe SARS-CoV-2 infections may continue shedding viral RNA for weeks or months after the acute infection, without having replication-competent virus, monitoring infection by molecular testing should be discouraged.

This study had several strengths and limitations. While having a larger sample size and more viral diversity would have been ideal, the study was not designed to compare diagnostic results across variant sub-types. Our analyses excluded several asymptomatic patients with acute SARS-CoV-2, who may be capable of transmitting infection.^51,52^ We also limited our nasal swab testing (i.e., RT-PCR, antigen, and culture) to nineteen days after enrollment and therefore may have overestimated estimates of time to negative test, since individuals who never tested negative during follow-up were excluded from the calculation. Strengths of the study were high retention rates for a population of symptomatic adults undergoing repeated invasive sampling procedures and consistency of trained medical assistant performing the swabs over time. The cohort was relatively young and healthy without immune altering conditions, which is more population representative than studies conducted among elderly or hospitalized populations.

In conclusion, we presented results from a first infection study of ambulatory adults with acute SARS-CoV-2 infection to describe and compare the longitudinal dynamics for viral viability (culture), viral load by RT-PCR, and viral S and N antigen quantification. The results underscore the importance of molecular RT-PCR testing as a highly sensitive diagnostic tool to determine the presence of viral RNA over the first four weeks of infection and also highlight the role for N antigen testing as an acceptable diagnostic test within two weeks of symptom onset given its stronger correlation to recovery of replication-competent virus and potential infectivity. Importantly, these findings indicate that public health guidance should encourage persons with acute SARS-CoV-2 infection to use N antigen testing (which are rather ubiquitous as rapid diagnostic tests), rather than the absence of symptoms, to safely discontinue an isolation period. These results may be used to strengthen infection control measures and reduce SARS-CoV-2 transmission to accelerate ending the COVID-19 pandemic.

## Data Availability

All data produced in the present work are contained in the manuscript

## Author contributions

PKD, GG, and MG conceived the study and acquired the funding. RD, PKD, MT, GG, and MG developed the study protocol. RD, JFM, ZM, RP, CW, AG, and PKD acquired the clinical data and specimens. RCI, ER, and DH provided important research infrastructure and sample processing. LH, ALG, MJM, BDG, JLC, ASB, and MJG conducted laboratory testing. MJB, EB, AM, RD, and PKD conducted the statistical analyses. PKD prepared the first manuscript draft. All authors provided critical feedback and approved of the final manuscript.

---

**Supplemental Table 1.** Average days since enrollment and symptom onset, and specimens tested, by participant visits.

|  | Visit #1 | Visit #2 | Visit #3 | Visit #4 | Visit #5<sup>a</sup> | Visit #6<sup>a</sup> | Total |
| --- | --- | --- | --- | --- | --- | --- | --- |
| **Participant attended visit** | 95 | 89 | 83 | 82 | 79 | 79 | **507** |
| Days since enrollment |  |  |  |  |  |  |  |
| Mean | 0.0 | 3.9 | 7.3 | 14.5 | 28.9 | 58.8 |  |
| Median [IQR] | 0 [0, 0] | 4 [3, 5] | 7 [7, 7] | 14 [14, 14] | 28 [28, 29] | 56 [56, 59] |  |
| Min, max | 0, 0 | 3, 6 | 7, 11 | 14, 19 | 27, 34 | 56, 82 |  |
| Days since symptom onset |  |  |  |  |  |  |  |
| Mean | 6.9 | 10.9 | 14.5 | 21.7 | 36.0 | 65.9 |  |
| Median [IQR] | 6 [5, 8] | 11 [9, 13] | 14 [12, 17] | 21 [19, 24] | 35 [34, 39] | 64 [62, 68] |  |
| Min, max | 1, 14 | 5, 18 | 8, 21 | 15, 29 | 29, 44 | 57, 87 |  |
| **Specimens Tested** |  |  |  |  |  |  |  |
| NP for viral load by RT-PCR | 95 | 88 | 82 | 82 | 0 | 2 | **349** |
| NP for viral culture | 83 | 81 | 69 | 74 | 0 | 2 | **309** |
| AN for nucleocapsid (N) antigen | 95 | 88 | 83 | 82 | 0 | 0 | **348** |
| AN for spike (S) antigen | 95 | 89 | 83 | 82 | 0 | 0 | **349** |
| Blood for Total anti-S antibodies | 93 | 89 | 83 | 82 | 79 | 78 | **504** |
| Blood for IgG anti-S antibodies | 93 | 89 | 83 | 82 | 79 | 78 | **504** |
| Viral sequencing (variants) | 46 | 18 | 3 | 3 | 0 | 0 | **81** |

AN=anterior nares; IQR=interquartile range; NP=nasopharyngeal.
<sup>a</sup>Nasal swabs were collected at visit #5 and #6 only for participants who reported any COVID-19-like or respiratory symptoms, and all tested were negative.

**Supplemental Table 2.** Diagnostic tests (S antigen, viral culture, N antigen, and RT-PCR for viral RNA) performed on samples collected at the first four visits. Median time (days) from symptom onset until negative test was determined for each test type, among participants with a negative test result during follow-up (see Figure 1).

| **N=95** | **Study exit before**<br>**negative test observed<sup>a</sup>**<br>N (%) | **End of follow-up before**<br>**negative test observed<sup>b</sup>**<br>N (%) | **Observed test-negatives**<br>N (%) | **Days since symptom onset of**<br>**observed test-negatives**<br>Median [IQR] |
| --- | --- | --- | --- | --- |
| S antigen | 4 (4) | 0 | 91 (96) | 9 [5] |
| Culture | 7 (7) | 0 | 88 (93) | 11 [4] |
| N antigen | 6 (6) | 1 (1) | 88 (93) | 13 [6] |
| RT-PCR | 12 (13) | 45 (47) | 38 (40) | >19<sup>c</sup> |

IQR=interquartile range.
<sup>a</sup>Of 95 sampled participants, some exited before a negative test could be observed (treated as non-informative and excluded from the median analysis).
<sup>b</sup>Other participants attended study visits but were positive at time of last collected nasal swabs for diagnostic testing (V4). If these individuals could have been observed longer, time to test negative is expected to be greater than the median estimated using observed test-negatives.
<sup>c</sup>Median [IQR] for RT-PCR could not be precisely estimated because more than half of participants were positive at all available sample times (>2-4 weeks after onset of symptoms).

**Supplemental Figure 1.**
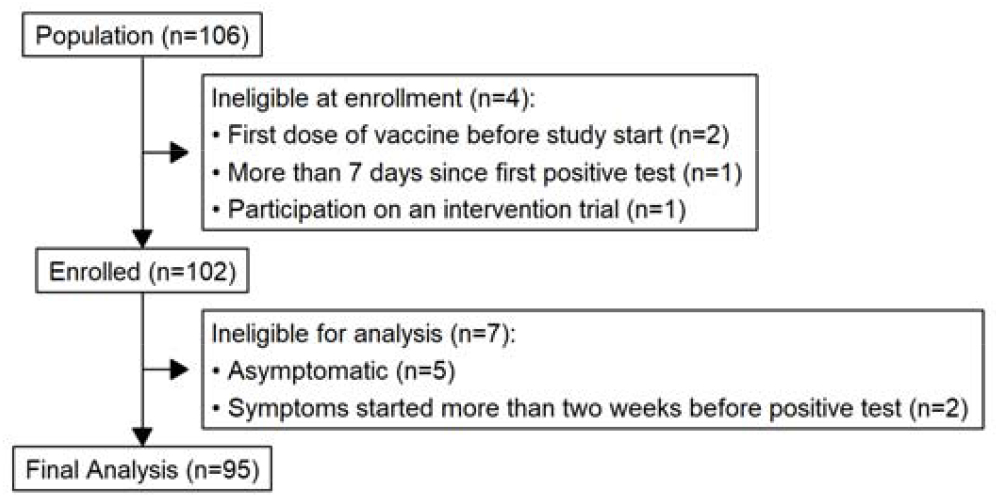
Consort diagram of study recruitment and enrollment. Among COVID-positive longitudinal study participants, of the 106 participants recruited, four were ineligible. Two people had received COVID-19 vaccination, one tested positive >7 days before enrollment, and one participated in a clinical trial). Seven participants were subsequently excluded from analyses: five because they remained asymptomatic, and two who were symptomatic >2 weeks prior to diagnostic testing.

**Supplemental Figure 2.**
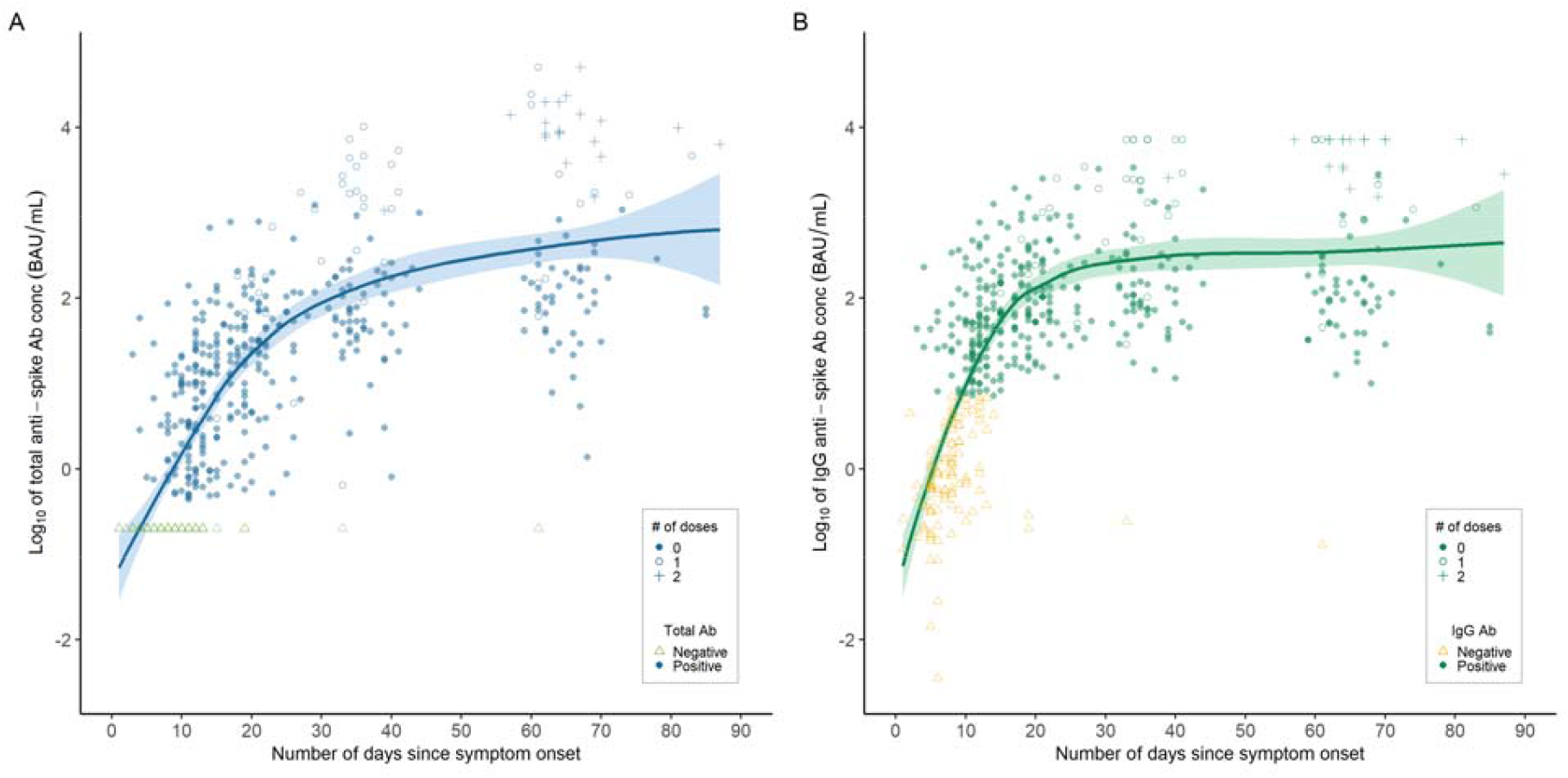
Trajectory of Total and IgG antibody titers, by days since symptom onset and with the inclusion of individuals who received COVID-19 vaccination during the study period. Average lines represent LOESS curves and shaded regions represent 95% confidence intervals. Color indicates qualitative result, and shape indicates vaccination status at the time of sample collection. **A**, Total anti-spike log_10_ mean antibody concentration (BAU/mL) tested by Roche Cobas e411 system (n=504). **B**, Anti-spike IgG log_10_ mean antibody concentration (BAU/mL) tested by Abbott AdviseDx platform (n=504).

**Supplemental Figure 3.**
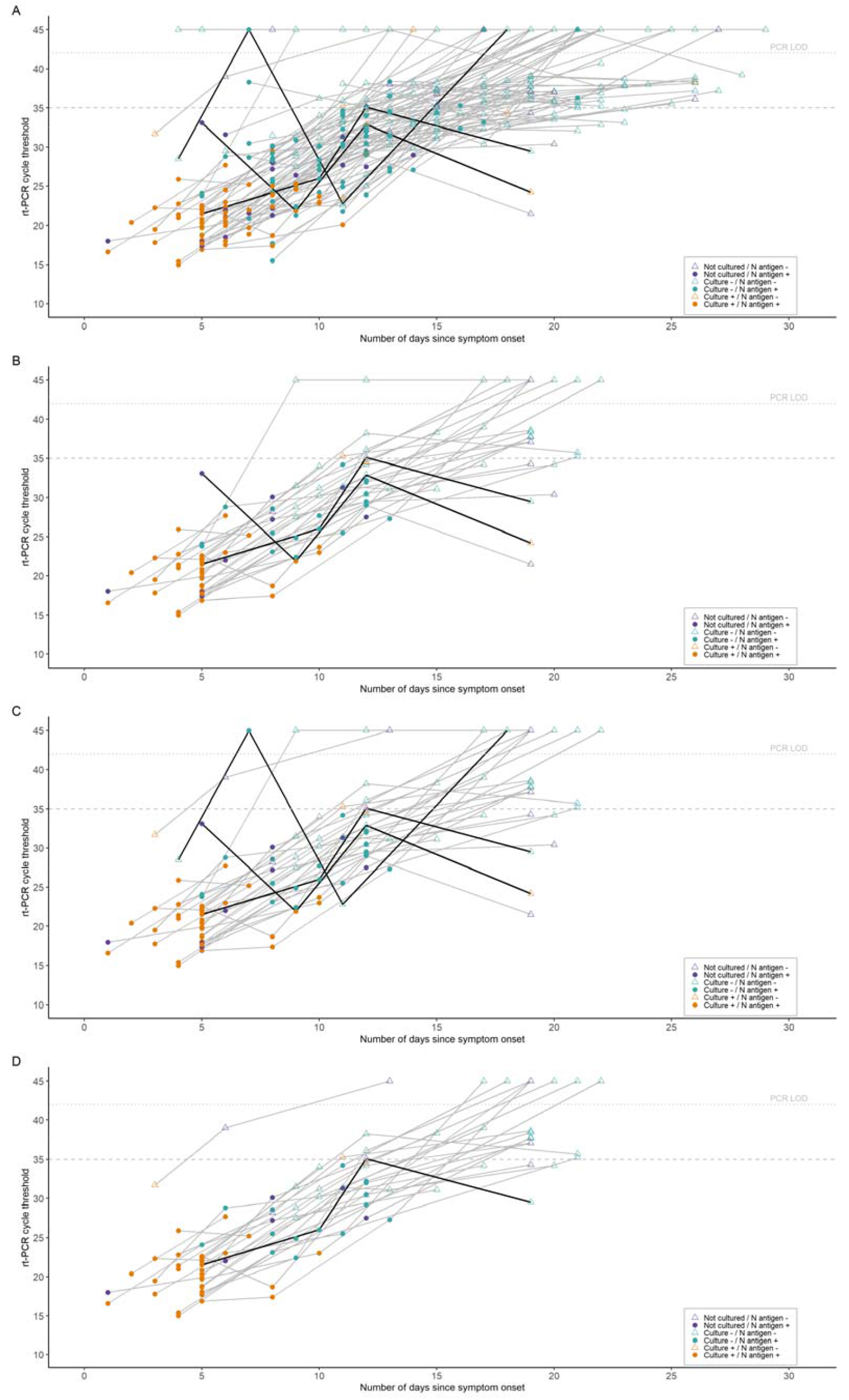
Individual trajectories for diagnostic test kinetics and infectivity during acute SARS-CoV-2 infection. Spaghetti plots show serial testing with one connected line for each individual participant indicating change in results over time. The x-axis shows days since symptom onset, and RT-PCR Ct value is indicated on y-axis. Viral culture result (positive/negative) is indicated by color (orange=viral growth; teal=no viral growth; purple=not cultured); matched N antigen test result is indicated by fill (hollow=negative, solid=positive). Horizontal lines present RT-PCR limit of detection (Ct=42.0) and Ct=35.0 for reference. **A**, All participants. **B**, Participants with positive N antigen test within five days of symptom onset. **C**, Participants with positive RT-PCR test within five days of symptom onset. **D**, Participants with positive viral culture result within five days of symptom onset.

## References

1. World Health Organization. WHO coronavirus (COVID-19) dashboard (2022); https://COVID19.who.int

2. Wu, S. L. et al. Substantial underestimation of SARS-CoV-2 infection in the United States. Nat. Commun. 11, 4507 (2020).

3. COVID-19 Excess Mortality Collaborators. Estimating excess mortality due to the COVID-19 pandemic: a systematic analysis of COVID-19-related mortality, 2020–21. Lancet Published Online March 10, 2022 https://doi.org/10.1016/S0140-6736(21)02796-3 (2022).

4. World Health Organization. Technical specifications for selection of essential in vitro diagnostics for SARS-CoV-2. Geneva; World Health Organization, 2021.

5. World Health Organization. Laboratory testing strategy recommendations for COVID-19: Interim Guidance. 21 March 2020. Ref: WHO/2019-nCoV/lab_testing/2020.1 https://apps.who.int/iris/bitstream/handle/10665/331509/WHO-COVID-19-lab_testing-2020.1-eng.pdf Note: “The role of rapid disposable tests for antigen detection for COVID-19 needs to be evaluated and is not currently recommended for clinical diagnosis pending more evidence on test performance and operational utility. WHO will update this guidance as more information laboratory tests for COVID-19 becomes available.”

6. Drain, P.K., et al. Diagnostic point-of-care tests in resource-limited settings. Lancet Infect Dis. 14, 239–49 (2014).

7. Yadav, H., Shah, D., Sayed, S., Horton, S., Schroeder, L.F. Availability of essential diagnostics in ten low-income and middle-income countries: results from national health facility surveys. Lancet Glob Health 9, e1553–60 (2021).

8. Drain, P. K. Rapid Diagnostic Testing for SARS-CoV-2. N Engl J Med. 386, 264–72 (2022).

9. World Health Organization. Antigen-detection in the diagnosis of SARS-CoV-2 infection: Interim guidance. WHO/2019-nCoV/Antigen_Detection/2021.1. https://www.who.int/publications/i/item/antigen-detection-in-the-diagnosis-of-sars-cov-2infection-using-rapid-immunoassays

10. Centers for Disease Control and Prevention. COVID-19: Testing for COVID-19 (2021). https://www.cdc.gov/coronavirus/2019-ncov/testing/index.html

11. Joint Research Center, European Commission. COVID-19 In Vitro Diagnostic Medical Devices, 2022. https://COVID-19-diagnostics.jrc.ec.europa.eu/devices?device_id=&manufacturer=&text_name=&marking=Yes&method=&rapid_diag=1&target_type=6&search_method=AND#form_content

12. Lee, J., Song, J.U., Shim, S.R. Comparing the diagnostic accuracy of rapid antigen detection tests to real time polymerase chain reaction in the diagnosis of SARS-CoV-2 infection: A systematic review and meta-analysis. J Clin Vir. 144, 104985 (2021).

13. Brümmer, L.E., et al. Accuracy of novel antigen rapid diagnostics for SARS-CoV-2: A living systematic review and metaanalysis. PLoS Med. 18, e1003735 (2021).

14. Hanson, K.E., et al. The Infectious Diseases Society of America guidelines on the diagnosis of COVID-19: Antigen testing. http://www.idsociety.org/COVID19guidelines/Ag.

15. Foundation for Innovative New Diagnostics. Test Directory, 2022. https://www.finddx.org/test-directory/.

16. Bekliz, M., et al. SARS-CoV-2 rapid diagnostic tests for emerging variants. Lancet Microbe 2, e351 (2021).

17. Mina, M.J., Andersen, K.G. COVID-19 testing: One size does not fit all. Science 371, 126–7 (2021).

18. Peeling, R.W., Olliaro, P.L., Boeras, D.I., Fongwen, N. Scaling up COVID-19 rapid antigen tests: promises and challenges. Lancet Infect Dis. 21, e290–5 (2021).

19. He, X., et al. Temporal dynamics in viral shedding and transmissibility of COVID-19. Nat Med. 26, 672–5 (2020).

20. Jones, T.C., et al. Estimating infectiousness throughout SARS-CoV-2 infection course. Science 373, eabi523 (2021).

21. Bullard, J., et al. Predicting infectious SARS-CoV-2 from diagnostic samples. Clin Infect Dis. 71, 2663–6 (2020).

22. Pekosz, A., et al. Antigen-based testing but not real-time polymerase chain reaction correlates with severe acute respiratory syndrome coronavirus 2 viral culture. Clin Infect Dis. 73, e2861–6 (2021).

23. Ricks, S. Quantifying the potential value of antigen-detection rapid diagnostic tests for COVID-19: a modelling analysis. BMC Med. 19, 75 (2021).

24. Drain, P.K., Garrett, N. SARS-CoV-2 pandemic expanding in sub-Saharan Africa considerations for COVID-19 in people living with HIV. EClinicalMedicine 22, 100342 (2020).

25. Peeling RW, Heymann DL, Teo Y-Y, Garcia PJ. Diagnostics for COVID-19: moving from pandemic response to control. Lancet 2022; 399 (10326): P757–68.

26. World Health Organization. Criteria for releasing COVID-19 patients from isolation; Scientific Brief. 17 June 2020. https://www.who.int/news-room/commentaries/detail/criteria-for-releasing-covid-19-patients-from-isolation

27. Centers for Disease Control and Prevention. COVID-19: Testing for COVID-19 (2022). https://www.cdc.gov/coronavirus/2019-ncov/your-health/quarantine-isolation.html

28. European Centre for Disease Control and Prevention. Guidance on ending the isolation period for peope with COVID-19, third update. 28 January 2022. https://www.ecdc.europa.eu/sites/default/files/documents/Guidance-for-discharge-and-ending-of-isolation-of-people-with-COVID-19-third-update.pdf

29. Center for Disease Control and Prevention. Symptoms of COVID-19 (2022). https://www.cdc.gov/coronavirus/2019-ncov/symptoms-testing/symptoms.html

30. World Health Organization. Coronavirus disease (COVID-19): Symptoms (2022). https://www.who.int/health-topics/coronavirus#tab=tab_3

31. O’Toole, A., et al. Assignment of epidemiological lineages in an emerging pandemic using the pangolin tool. Virus Evolution. 7, veab064 (2021).

32. Bachman, C.M., et al. Clinical validation of an open-access SARS-COV-2 antigen detection lateral flow assay, compared to commercially available assays. PLoS ONE 16, e0256352 (2021).

33. Debad, J., Glezer, E., Wohlstadter, J., Sigal, G. Clinical and biological applications of ECL. Electrogenerated chemiluminescence. Marcel Dekker, New York, NY; 2004: 43–78.

34. US Centers for Disease Control and Prevention. Viral Transport Medium, 2021. https://www.cdc.gov/coronavirus/2019-ncov/downloads/Viral-Transport-Medium.pdf.

35. Montaño, M.A., et al. Performance of anterior nares and tongue swabs for nucleic acid, nucleocapsid, and spike antigen testing for detecting SARS-CoV-2 against nasopharyngeal PCR and viral culture. Int J Infect Dis. 117, 287–294 (2022).

36. Cantera, J.L., et al. Screening Antibodies Raised against the Spike Glycoprotein of SARS-CoV-2 to Support the Development of Rapid Antigen Assays. ACS Omega. 6, 20139–20148 (2021).

37. Perkmann, T., et al. Anti-Spike Protein Assays to Determine SARS-CoV-2 Antibody Levels: a Head-to-Head Comparison of Five Quantitative Assays. Microbiol Spectr. 9, e00247–21 (2021).

38. Mina, M.J., Parker, R., Larremore, D.B. Rethinking COVID-19 test sensitivity–a strategy for containment. N Engl J Med. 383, e120 (2020).

39. Killingley, B., et al. Safety, tolerability and viral kinetics during SARS-CoV-2 human challenge in young adults. Nat. Med. (2022). https://doi.org/10.1038/s41591-022-01780-9

40. The COVID-19 Investigation Team. Clinical and virologic characteristics of the first 12 patients with coronavirus disease 2019 (COVID-19) in the United States. Nat Med 26, 861–868 (2020).

41. Weissz, A., Jellingsø, M., Sommer, M.O.A. Spatial and temporal dynamics of SARS-CoV-2 in COVID-19 patients: A systematic review and meta-analysis. EBioMedicine 58, 102916 (2020).

42. Hartman, W.R., Hess, A.S., Connor, J.P. Persistent viral RNA shedding after COVID-19 symptom resolution in older convalescent plasma donors. Transfusion, 60, 2189–2191 (2020).

43. Liu, Y., et al. Viral dynamics in mild and severe cases of COVID-19. Lancet Infect Dis. 20, 656–7 (2020).

44. Zhou, B., She, J., Wang, Y., Ma, X. The duration of viral shedding of discharged patients with severe COVID-19. Clin Infect Dis. 71, 2240–2 (2020).

45. Zou, L., Ruan, F., Huang, M., Liang, L., Huang, H., Hong, Z. SARS-CoV-2 viral load in upper respiratory specimens of infected patients. N Engl J Med. 382, 1177–1179 (2020).

46. Lee, S., et al. Clinical course and molecular viral shedding among asymptomatic and symptomatic patients with SARS-CoV-2 infection in a community treatment center in the Republic of Korea. JAMA Int Med. 180, 1447–52 (2020).

47. Wölfel, R., et al. Virological assessment of hospitalized patients with COVID-2019. Nature 581, 465–9 (2020).

48. Seow, J, et al. Longitudinal evaluation and decline of antibody responses in SARS-CoV-2 infection. Nature Microbiol. 5, 1598–607 (2020).

49. Mizrahi, B., et al. Longitudinal symptom dynamics of COVID-19 infection. Nature Commun. 11, 6208 (2020).

50. Nielsen, K.J., et al. Day-by-day symptoms following positive and negative PCR tests for SARS-CoV-2 in non-hospitalized healthcare workers: A 90-day follow-up study. Int J Infect Dis. 108, 382–90 (2021).

51. Buitrago-Garcia, D., et al. Occurrence and transmission potential of asymptomatic and pre-symptomatic SARS-CoV-2 infections: A living systematic review and meta-analysis. PLoS Med. 17, e1003346 (2020).

52. Qiu, X, et al. The role of asymptomatic and pre-symptomatic infection in SARS-CoV-2 transmission—a living systematic review. Clin Microbiol Infect. 27, 511–9 (2021).

